# The prevalence of common mental disorders among health care professionals during the COVID-19 pandemic at a tertiary Hospital in East Africa

**DOI:** 10.1101/2020.10.29.20222430

**Authors:** Hailu Abera Mulatu, Muluken Tesfaye, Esubalew Woldeyes, Tola Bayisa, Henok Fesseha, Rodas Asrat

## Abstract

**Background:** Coronavirus disease 2019 (COVID-19) has resulted in unprecedented morbidity, mortality, and health system crisis leading to a significant psychological distress on healthcare workers (HCWs). The study aimed to determine the prevalence of symptoms of common mental disorders among HCWs during the COVID-19 pandemic at St. Paul’s Hospital, Ethiopia.

**Methods:** A self-administered cross-sectional study was conducted to collect socio-demographic information and symptoms of mental disorders using validated measurement tools. Accordingly, PHQ-9, GAD-7, ISI, and IES-R were used to assess the presence of symptoms of depression, anxiety, insomnia, and distress, respectively. Chi-square test, non-parametric, and logistic regression analysis were used to detect risk factors for common mental disorders.

**Results:** A total of 420 healthcare workers participated in the survey. The prevalence of depression, anxiety, insomnia, and psychological distress was 20.2%, 21.9%, 12.4%, and 15.5% respectively. Frontline HCWs had higher scores of mental health symptoms than other health care workers. Logistic regression analysis showed that being married was associated with a high level of depression. Working in a frontline position was an independent factor associated with a high-level depression, anxiety, and psychological distress.

**Limitations:** It is a single-centre cross-sectional study and the findings may not be generalizable or reveal causality.

## 1. Introduction

Coronavirus disease 2019 (COVID-19) is a mild to severe respiratory illness that is caused by a coronavirus named severe acute respiratory syndrome coronavirus 2 (SARS COV2). It was first recognized in China in late December 2019 as an unknown respiratory disease outbreak. Due to its high risk of contagiousness and human-to-human transmission, it has reached global pandemic level in a very short time (Guan *et al*., 2020; C. Lai *et al*., 2020). As of September 18, 2020, WHO has reported 30,055,710 confirmed cases of COVID-19, and 943,433 deaths globally.(WHO, 2020) On the same day, the Ethiopian Ministry of Health has also reported 67,515 total confirmed cases and 1,072 deaths in the country (MOH, 2020). Healthcare workers (HCWs) dealing with COVID-19 are under increased psychological pressure, and experience high rates of psychiatric morbidity, resembling the situation during the previous severe acute respiratory syndrome (SARS) and influenza epidemics (Li *et al*., 2020). Depression, anxiety, insomnia, and psychological distress are the common mental health disorders that occur during such a dramatic global health crisis (Liu *et al*., 2020; Pappa *et al*., 2020). A recent study among 1257 healthcare professionals in a tertiary hospital in China, revealed a high prevalence of mental health symptoms among HCWs. Overall, 50.4%, 44.6%, 34.0%, and 71.5% of health workers reported symptoms of depression, anxiety, insomnia, and distress, respectively. Nurses, females, and frontline HCWs were affected more than others with these mental disorders (Bohlken *et al*., 2020).

To date, studies on the psychological impact of COVID-19 on HCWs are limited in Sub Saharan Africa. The aims of the current study were to determine the prevalence of depression, anxiety, insomnia, and psychological distress among HCWs and associated factors during the COVID-19 pandemic at St. Paul’s Tertiary Hospital, Addis Ababa, Ethiopia.

## 2. Materials and Methods

### 2.1 Study Setting and design

The study was conducted at St. Paul’s Hospital, the second-largest public hospital in Ethiopia located in Addis Ababa. There are 2308 HCWs involved in clinical care. The health professionals comprise of 323 specialists, 432 general practitioners, 1208 nurses, 135 midwives, and 210 laboratory and pharmacy professionals. The hospital has an inpatient capacity of more than 700 beds and serves an average of 1200 emergency and outpatient clients daily. After the first few cases of COVID-19 were reported in March 2020 in Ethiopia, the hospital has dedicated 350 ward and 14 ICU beds to treating only these cases.

The study was a cross-sectional, hospital-based survey that was conducted to assess the prevalence of depression, anxiety, insomnia, and mental distress among healthcare workers at St. Paul’s Hospital during the COVID-19 pandemic from August 1^st^, 2020 up to August 30, 2020.

The sample size was determined based on a single population formula for a finite population of 2308 HCWs with a 95% CI, 5% margin of error, and taking the highest of the prevalence of depression, anxiety, insomnia, and distress of 50.4% (Lai *et al*., 2020; Sofia *et al*., 2020; Zhu *et al*., 2020), 44.6% (Lai *et al*., 2020; Sofia *et al*., 2020; Zhu *et al*., 2020), 34.3% (Lai *et al*., 2020; Sofia *et al*., 2020;), and 71.5% (Lai *et al*., 2020; Zhu *et al*., 2020) respectively from similar studies. By considering a 10% non-response rate and applying a sample correction formula, a total sample size of 272 HCWs will be included in the study. To allow for subgroup analysis, we amplified the sample size by 50% with a goal of at least 430 completed questionnaires from participants. Healthcare workers working in either in-patient or out-patient units or COVID or non-COVID units of the Hospital were included. Healthcare workers who had previous mental illnesses, not involved in the diagnosis or treatment of patients, and those who did not give consent were excluded from the study.

### 2.2 Operational Definitions

**Healthcare worker (HCW):** health professionals (medical doctors, nurses, midwives, laboratory professionals and pharmacists) who have close contact with patients in the diagnostic and treatment units of Saint Paul’s Hospital Millennium Medical College.

**Frontline HCWs:** participants who are directly engaged in clinical activities of diagnosing, treating, or providing nursing care to patients with confirmed COVID-19.

**Non-firstline (Second-line) HCWs:** participants who are not directly engaged in clinical activities of diagnosing, treating, or providing nursing care to patients with confirmed COVID-19 but could be indirectly exposed while involved in the care of other patients who might be in a pre-symptomatic stage of COVID-19.

### 2.3 Sampling Method and Sampling Procedure

Healthcare workers of both sexes working in both out-patient and in-patient units of St. Paul’s Hospital were illegible for the study. Participants were recruited using a simple random sampling method from their alphabetical list prepared by the human resource development directorate after first stratifying them into four groups (doctors, nurses/ midwives, the laboratory professionals, and pharmacy professionals) using probability proportional to size method. All volunteered study subjects completed a self-administered questionnaire that was used to assess for symptoms of depression, anxiety, insomnia, and distress.

### 2.4 Data collection tools and procedures

Data were collected by trained residents and nurses.The data collectors categorized the participants by asking whether they were directly engaged in clinical activities of diagnosing, treating, or providing nursing care to patients with confirmed COVID-19. Those who responded yes were defined as frontline workers, and those who answered no were defined as non-frontline (second-line) workers. Data were collected using a pretested and self-administered questionnaire to assess for symptoms of depression, anxiety, insomnia, and distress using Amharic versions of validated measurement tools (Gelaye, *et al*., 2014; Hanlon *et al*., 2015). Accordingly, the 9-item Patient Health Questionnaire (PHQ-9; range, 0-27), the 7-item Generalized Anxiety Disorder (GAD-7) scale (range, 0-21), the 7-item Insomnia Severity Index (ISI; range, 0-28), and the 22-item Impact of Event Scale-Revised (IES-R; range, 0-88) were used to assess the severity of symptoms of depression, anxiety, insomnia, and distress, respectively. The total scores of these measurement tools were interpreted as follows: PHQ-9, normal (0-4), mild (5-9), moderate (10-14), and severe (15-21) depression; GAD-7, normal (0-4), mild (5-9), moderate (10-14), and severe (15-21) anxiety; ISI, normal (0-7), sub-threshold (8-14), moderate (15-21), and severe (22-28) insomnia; and IES-R, normal (0-8), mild (9-25), moderate (26-43), and severe (44-88) distress. These categories were based on values established in the literature (Gelaye B *et al*., 2014; Hanlon *et al*., 2015; Greenberg *et al*., 2020). The cut-off score for detecting severe symptoms of depression, anxiety, insomnia, and distress were 10 (Johnson *et al*., 2019), 9 (Ong, & Suh, 2013), 15 (Ong, J. C., & Suh, 2013; Johnson *et al*., 2019), and 33 (Creamer *et al*., 2003), respectively. Participants who scored greater than or equals to the cut-off threshold were characterized as having severe symptoms. The selected 4 questionnaires had good internal consistency with Cronbach’s α coefficients of more than 0.80. Demographic data were self-reported by the participants, including educational level (graduate, post-graduate), profession (physician, nurse/midwife, the laboratory professional or pharmacy professional), sex (male or female), age (18-25, 26-30, 31-40, or >40 years), marital status, monthly net salary, educational level (undergraduate or postgraduate), and technical title (junior or senior). The different technical titles of respondents refer to the professional titles certificated by the Ethiopian Food, Medicine and Health Care Administration, and Control Authority (EFMHACA). All investigators supervised the data collection process.

### 2.5 Data processing and analysis

Each completed questionnaire was entered into EPI data version 7, cleaned, and exported into SPSS version 23 for statistical analysis. All the variables were categorical and presented as numbers and percentages. The original scores of the 4 measurement tools were continuous, not normally distributed, and thus presented as a median and interquartile range (IQR). A nonparametric Mann-Whitney U test and Kruskal-Wallis H test were applied to compare the severity of each symptom between two or more groups. The ranked data, which were derived from the counts of each level for symptoms of depression, anxiety, insomnia, and distress were presented as numbers and percentages. To determine potential risk factors for symptoms of depression, anxiety, insomnia, and distress in participants, a multivariable logistic regression analysis was performed, and the associations between risk factors and outcomes were presented as odds ratios (ORs) and 95% CIs, after adjustment for confounders, including age, sex, marital status, profession, educational level, technical rank, monthly salary, work experience, and working position (frontline or non-frontline). Finally, a p-value <0.05 in a multivariable model was considered as significant.

### 2.6 Ethical approval

This study was approved by the Institutional Review Board of St. Paul’s Hospital Millennium Medical College. Written informed consent was obtained from all participants. Each participant’s information was collected by residents and nurses using an anonymous pre-coded structured questionnaire that was assigned to each selected HCW. The code was blinded to both data collectors and data entry clerks. The information was kept confidential among the investigators. A participant who had severe symptoms of mental disorder was traced by a psychiatrist (one of the investigators) and advised on further evaluation and treatment. Those who volunteered were linked to our psychiatric clinic.

## 3. Results

### 3.1 Socio-demographic characteristics of study participants

A total of 420 participants were included in the study making a response rate of 97.7%. Of these, 115 (27.4%) participants were physicians, 237 (56.4%) were nurses and midwives, 40 (9.5%) were laboratory professionals and 28 (6.7%) were pharmacy professionals. The response rates for physicians, nurses or midwives, laboratory professionals, and pharmacy professionals were 92%, 100%, 100%, and 100% respectively. The mean age of the participants was 28±5.4 [range: 20-57] years. Among the participants, 212 (50.5%) were in the age range of 26-30 years. Two-hundred-forty-six (58.6%) participants were men and 296 (70.5%) were unmarried. The qualification of participants indicated that 295 (70.2%) had undergraduate level and 125 (29.8%) had a postgraduate level of education. Regarding their technical title, 237 (56.4%) were junior and 183 (43.6%) were senior staff in their professional career. The mean monthly salary of respondents was 7527± 3819 Ethiopian Birr (ETB) and 213 (50.7%) respondents earn between 5000-10,000 ETB. The majority (70.5%) of respondents had a work experience of fewer than five years and 296 (70.5%) were frontline healthcare professionals. A significant proportion of respondents working as frontline were aged between 26-30 years, unmarried, and nurses or midwives. They had an undergraduate level of education and work experience of fewer than five years (Table 1).

**Table 1.** Socio-demographic characteristics of participants, St. Paul’s Hospital, 2020.

| Characteristic |  | Number (%) |  |  | p-value |
| --- | --- | --- | --- | --- | --- |
|  |  | Overall<br>420 | Frontline<br>296 | Non-frontline<br>124 |  |
| Age, year | 18-25 | 121 (28.8) | 84 (28.4) | 37 (29.8) | <0.001 |
|  | 26-30 | 212 (50.5) | 165 (55.7) | 47 (37.9) |  |
|  | 31-40 | 75 (17.9) | 44 (14.9) | 31 (25.0) |  |
|  | >40 | 12 (2.9) | 3 (1.0) | 9 (7.3) |  |
| Sex | Male | 246 (58.6) | 176 (59.5) | 70 (56.5) | 0.56 |
|  | Female | 174 (41.4) | 120 (40.5) | 54 (43.5) |  |
| Marital status | Unmarried | 296 (70.5) | 223 (75.3) | 73 (58.9) | 0.001 |
|  | Ever married* | 124 (29.5) | 73 (24.7) | 51 (41.1) |  |
| Profession | Physician | 115 (27.4) | 73 (24.7) | 42 (33.9) | <0.001 |
|  | Nurse/Midwife | 237 (56.4) | 177 (59.8) | 60 (48.4) |  |
|  | Laboratory profession | 40 (9.5) | 34 (11.5) | 6 (4.8) |  |
|  | Pharmacy profession | 28 (6.7) | 12 (4.0) | 16 (12.9) |  |
| Level of Education | undergraduate | 295 (70.2) | 218 (73.6) | 77 (62.1) | 0.02 |
|  | Postgraduate | 125 (29.8) | 78 (26.4) | 47 (37.9) |  |
| Technical title | Junior | 237 (56.4) | 171 (57.8) | 66 (53.2) | 0.45 |
|  | Senior | 183 (43.6) | 125 (42.2) | 58 (46.8) |  |
| Monthly Salary, ETB | <5,000 | 110 (26.2) | 87 (29.4) | 23 (18.5) | 0.01 |
|  | 5,000-10,000 | 213 (50.7) | 151 (51.0) | 62 (50.0) |  |
|  | >10,000 | 97 (23.1) | 58 (19.6) | 39 (31.5) |  |
| Work experience, y | <5 | 301 (71.7) | 217 (73.3) | 84 (67.7) | 0.02 |
|  | 5-10 | 93 (22.1) | 67 (22.6) | 26 (21.0) |  |
|  | >10 | 26 (6.2) | 12 (4.1) | 14 (11.3) |  |

ETB, Ethiopian Birr; SD, standard deviation; \*Ever married included widowed and divorced participants

### 3.2 The prevalence of mental disorders

The prevalence of depression, anxiety, insomnia, and psychological distress was found to be 20.2%, 21.9%, 12.4%, and 15.5% respectively. The median (IQR) scores on the PHQ-9 for depression, the GAD-7 for anxiety, the ISI for insomnia, and the IES-R for distress for all respondents were 4.0 (1.0-9.0), 3.0 (0-8.0), 4.0 (1.0-9.0), and 11.0 (4.0-25.0), respectively. Symptoms of depression, anxiety, insomnia, and distress were reported in higher proportion of frontline healthcare workers (i.e. depression among frontline vs non-frontline : 70 [23.6%] vs 15 [17.6%]; P<0.001; anxiety among frontline vs non-frontline: 75[25.3%] vs 17 [13.7%]; P = .003; insomnia among frontline vs non-frontline: 43[14.5%] vs 9 [7.3%]; P = .003; and distress among frontline vs non-frontline: 60 [20.3%] vs 5 [4.0%]; P < 0.001) (Table 2).

**Table 2.** Number of cases and scores of depression, anxiety, insomnia, and distress measurements in total cohort and subgroups among HCWs at St. Paul’s Hospital, 2020.

| Scale | Total cases (%) | Total Score, median (IQR) | Working position |  |  |
| --- | --- | --- | --- | --- | --- |
|  |  |  | Score, median (IQR) |  | P -value |
|  |  |  | Frontline | Non-frontline |  |
| PHQ-9, depression symptoms | 85 (20.2) | 4.0 (1.0-9.0) | 5.0 (2.0-9.0) | 2.0 (0-6.0) | **<0.001** |
| GAD-7, anxiety symptoms | 92 (21.9) | 3.0 (0-8.0) | 4.0 (0-9.0) | 2.5 (0-5.0) | **0.003** |
| ISI, insomnia symptoms | 52 (12.4) | 4.0 (1.0-9.0) | 5.0 (1.0-10.0) | 4.0 (1.0-7.0) | **0.003** |
| IES-R, distress symptoms | 65(15.5) | 11.0 (4.0-25.0) | 13.0 (4.25-29.0) | 8.0 (3.0-16.0) | **<0.001** |

Abbreviations: GAD-7, 7-item Generalized Anxiety Disorder; IES-R, 22-item Impact of Event Scale-Revised; ISI, 7-item Insomnia Severity Index; PHQ-9, 9-item Patient Health Questionnaire, IQR, interquartile range

### 3.3 Severity of symptoms of mental disorders and associated factors

Respondents were categorized based on the severity of symptoms of depression, anxiety, insomnia, and psychological distress. Moderate symptoms of depression, anxiety insomnia, and distress were reported in 57 (13.6%), 49 (11.7%), 43 (10.2%), and 71 (16.9%) respondents. Severe symptoms of depression, anxiety insomnia, and distress were also reported in 28 (6.7%), 24 (5.7%), 9 (2.1%), and 30 (7.1%) respondents. Respondents working in frontline reported experiencing moderate to severe symptoms of depression (i.e. moderate to severe depression among frontline vs non-frontline: 70 [23.6%] vs 15 [12.1%]). Moderate to severe symptoms of anxiety, insomnia, and distress were also more common among frontline HCWs (Table 3).

**Table 3.**
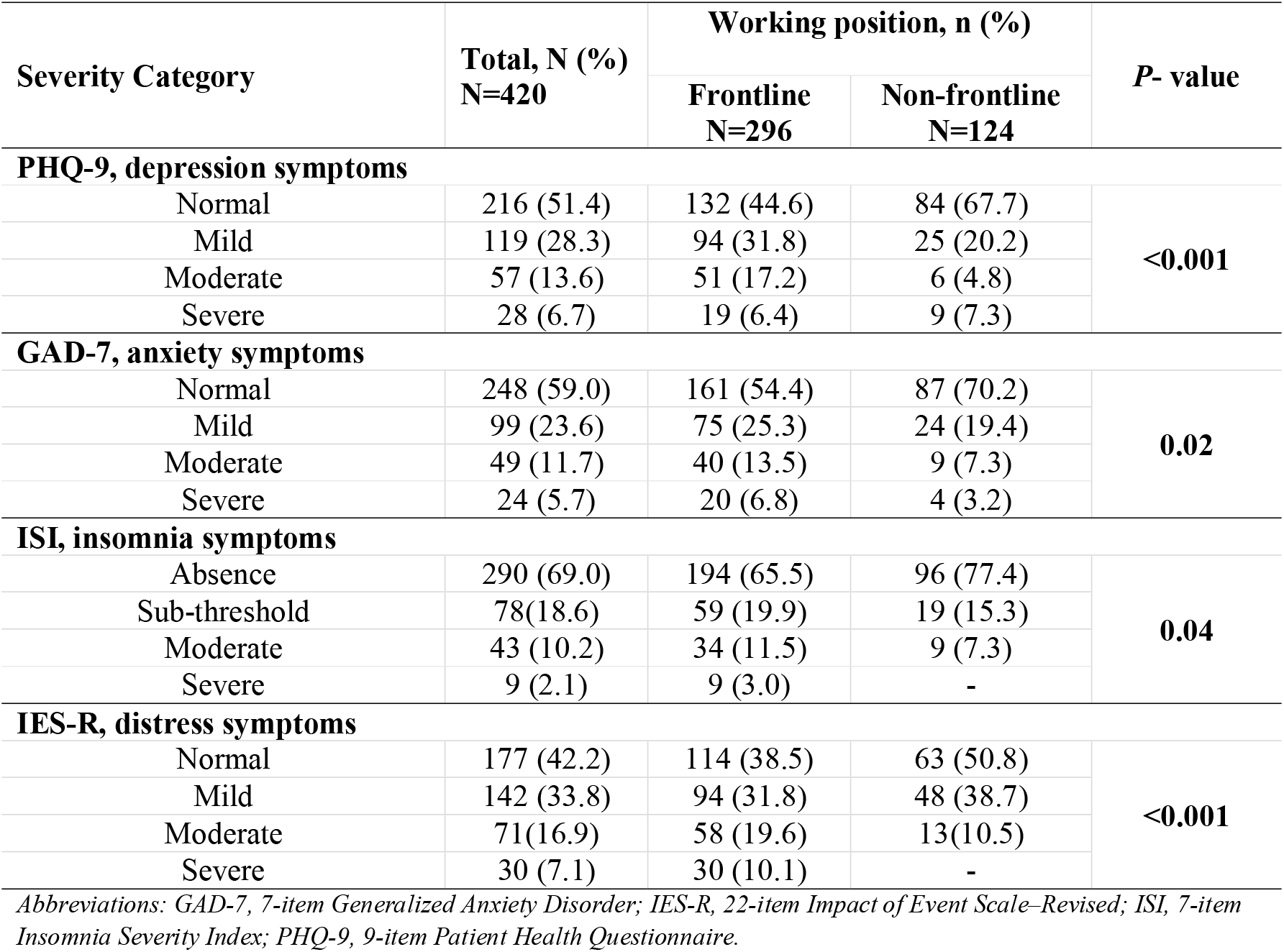
Severity categories of depression, anxiety, stress, insomnia, and psychological distress in total cohort and subgroups, St. Paul’s Hospital, 2020.

| Severity Category | Total, N (%)<br>N=420 | Working position, n (%) |  | P- value |
| --- | --- | --- | --- | --- |
|  |  | Frontline<br>N=296 | Non-frontline<br>N=124 |  |
| PHQ-9, depression symptoms |  |  |  |  |
| Normal | 216 (51.4) | 132 (44.6) | 84 (67.7) | <0.001 |
| Mild | 119 (28.3) | 94 (31.8) | 25 (20.2) |  |
| Moderate | 57 (13.6) | 51 (17.2) | 6 (4.8) |  |
| Severe | 28 (6.7) | 19 (6.4) | 9 (7.3) |  |
| GAD-7, anxiety symptoms |  |  |  |  |
| Normal | 248 (59.0) | 161 (54.4) | 87 (70.2) | 0.02 |
| Mild | 99 (23.6) | 75 (25.3) | 24 (19.4) |  |
| Moderate | 49 (11.7) | 40 (13.5) | 9 (7.3) |  |
| Severe | 24 (5.7) | 20 (6.8) | 4 (3.2) |  |
| ISI, insomnia symptoms |  |  |  |  |
| Absence | 290 (69.0) | 194 (65.5) | 96 (77.4) | 0.04 |
| Sub-threshold | 78(18.6) | 59 (19.9) | 19 (15.3) |  |
| Moderate | 43 (10.2) | 34 (11.5) | 9 (7.3) |  |
| Severe | 9 (2.1) | 9 (3.0) | - |  |
| IES-R, distress symptoms |  |  |  |  |
| Normal | 177 (42.2) | 114 (38.5) | 63 (50.8) | <0.001 |
| Mild | 142 (33.8) | 94 (31.8) | 48 (38.7) |  |
| Moderate | 71(16.9) | 58 (19.6) | 13(10.5) |  |
| Severe | 30 (7.1) | 30 (10.1) | - |  |

Abbreviations: GAD-7, 7-item Generalized Anxiety Disorder; IES-R, 22-item Impact of Event Scale-Revised; ISI, 7-item Insomnia Severity Index; PHQ-9, 9-item Patient Health Questionnaire.

### 3.4 Perception of threat of COVID-19 and effects of psychological protective measures

Regarding perception of threat of COVID-19, 252 (60%) respondents feel that they might have history of exposure to COVID-19, 80 (19%) had thought of resigning from work, 197 (46.9%) were worried of life-threatening situation, 283 (67.4%) feel that families and friends have avoided contact with them due to their work, and 362 (86.2%) were worried about their family members being infected by COVID-19.

Thought of resigning from work due to COVID-19 was reported in significantly higher proportion of respondents with symptoms of depression, anxiety, insomnia, and psychological distress (p-values <0.01). Worrying of life-threatening situation once infected was reported in a significant proportion of respondents with symptoms of depression (OR, 2.2; 95% CI, 1.2-3.8; p = 0.007). The study also assessed whether or not the respondents were satisfied with the psychological protective measures implemented at St. Paul’s Hospital. Respondents who were satisfied with the work-shift arrangement, logistic support, and accommodation suffered less from insomnia and psychological distress than those who were not satisfied with these protective measures (p≤0.01) (Table 4).

**Table 4.** Perceptions of threat of the COVID-19 and effects of psychological protective measures among HCWs dealing with the COVID-19 at St. Paul’s Hospital, 2020.

| Variable |  | No. of severe cases/ No. of total cases (%) | OR (95% CI) | P value |
| --- | --- | --- | --- | --- |
| PHQ-9, depression symptoms |  |  |  |  |
| Thought of resigning from work due to COVID-19 | Yes | 29/80 (36.3) | 2.3 (1.3-4.0) | 0.005 |
|  | No | 56/340 (16.5) | 1 [Reference] |  |
| Worried about a life-threatening situation once infected | Yes | 54/197 (27.4) | 1.8 (1.0-3.0) | 0.03 |
|  | No | 31/223 (13.9) | 1 [Reference] |  |
| GAD-7, anxiety symptoms |  |  |  |  |
| Thought of resigning from work due to COVID-19 | Yes | 28/80 (35.0) | 2.2 (1.2-3.8) | 0.007 |
|  | No | 64/340 (18.8) | 1 [Reference] |  |
| ISI, insomnia symptoms |  |  |  |  |
| Thought of resigning from work due to COVID-19 | Yes | 26/80 (32.5) | 5.0 (2.6-9.6) | <0.001 |
|  | No | 26/340 (7.6) | 1 [Reference] |  |
| Satisfied with work-shift arrangement | Yes | 13/206 (6.3) | 0.4 (0.2-0.8) | 0.005 |
|  | No | 39/214 (18.2) | 1 [Reference] |  |
| Satisfied with logistic support and accommodation | Yes | 2/109 (1.8) | 0.1 (0.03-0.5) | 0.003 |
|  | No | 50/311 (16.1) | 1 [Reference] |  |
| IES-R, distress symptoms |  |  |  |  |
| Thought of resigning from work due to COVID-19 | Yes | 35/80 (43.8) | 6.5 (3.5-12.0) | <0.001 |
|  | No | 30/340 (8.8) | 1 [Reference] |  |
| Satisfied with work-shift arrangement | Yes | 19/206 (9.2) | 0.45 (0.2-0.8) | 0.008 |
|  | No | 46/214 (21.5) | 1 [Reference] |  |
| Satisfied with logistic support and comfortable accommodation | Yes | 4/109 (3.7) | 0.2 (0.06-0.5) | 0.002 |
|  | No | 61/311 (19.6) | 1 [Reference] |  |

Abbreviation: GAD-7, 7-item Generalized Anxiety Disorder; IES-R, 22-item Impact of Event Scale-Revised; ISI, 7-item Insomnia Severity Index; OR, odds ratio; PHQ-9, 9-item Patient Health Questionnaire.

### 3.5 Factors associated with symptoms of mental health disorders

A multivariable logistic regression analysis was conducted to determine the demographic and relevant contextual factors that are associated with mental health illnesses. In the adjusted logistic regression analysis, several factors were independently associated with depression (PHQ-9 score ≥ 10), anxiety (GAD-7 score ≥ 9), insomnia (ISI score ≥ 15), and psychological distress (IES-R score ≥ 33). Factors that were independently associated with a higher risk of depression included being married (OR, 2.5; 95% CI, 1.4-4.6; p=0.003) and working in frontline position (OR, 2.4; 95% CI, 1.2-4.5; p=0.009). Working in frontline position is also associated with more severe symptoms of anxiety (OR, 2.1; 95% CI, 1.1-3.9; p=0.02) and psychological distress (OR, 5.9; 95% CI, 2.2-15.5; p<0.001). Working as a doctor was associated with a lower risk of experiencing anxiety (OR, 0.12; 95% CI, 0.03-0.51; p = 0.004) and insomnia (OR, 0.08; 95% CI, 0.02-0.59; p=0.013) than working as a laboratory or pharmacy professional (Table 5).

**Table 5.** Factors associated with mental health symptoms in multivariable logistic regression analysis.

| Variable |  | No. of severe cases/ No. of total cases (%) | Adjusted OR (95% CI)<sup>a</sup> | P value<sup>b</sup> |  |
| --- | --- | --- | --- | --- | --- |
|  |  |  |  | Category | Overall |
| PHQ-9, depression symptoms |  |  |  |  |  |
| Marital status | Ever-married | 32/124 (25.8) | 2.5 (1.4-4.6) | 0.003 | 0.003 |
|  | Unmarried | 53/296 (17.9) | 1 [Reference] | NA |  |
| Working position | Frontline | 70/296 (23.6) | 2.4 (1.2-4.5) | 0.009 | 0.006 |
|  | Non-frontline | 15/124 (12.1) | 1 [Reference] | NA |  |
| GAD-7, anxiety symptoms |  |  |  |  |  |
| Profession | Doctor | 7/115 (6.1) | 0.12 (0.03-0.51) | 0.004 | 0.01 |
|  | Nurse or midwife | 63/237 (26.6) | 0.66 (0.36-1.22) | 0.19 |  |
|  | Laboratory or pharmacy professional | 22/68 (32.4) | 1 [Reference] | NA |  |
| Working position | Frontline | 75/296 (25.3) | 2.1 (1.1-3.9) | 0.02 | 0.01 |
|  | Non-frontline | 17/124 (13.7) | 1 [Reference] | NA |  |
| ISI, insomnia symptoms |  |  |  |  |  |
| Profession | Doctor | 3/115 (2.6) | 0.08 (0.02-0.59) | 0.013 | 0.011 |
|  | Nurse or midwife | 33/237 (13.9) | 0.47 (0.23-0.96) | 0.04 |  |
|  | Laboratory or pharmacy professional | 16/68 (23.5) | 1 [Reference] | NA |  |
| IES-R, distress symptoms |  |  |  |  |  |
| Working position | Frontline | 60/296 (20.3) | 5.9 (2.2-15.5) | <0.001 | <0.001 |
|  | Non-frontline | 5/124 (4.0) | 1 [Reference] | NA |  |

Abbreviation: GAD-7, 7-item Generalized Anxiety Disorder; IES-R, 22-item Impact of Event Scale-Revised; ISI, 7-item Insomnia Severity Index; OR, odds ratio; PHQ-9, 9-item Patient Health Questionnaire; NA, not applicable; <sup>a</sup> Adjusted for age, sex, marital status, profession, educational level, technical title, salary, work experience, and working position, when appropriate; <sup>b</sup> Category refers to the P value for each category vs the reference, while overall refers to the results of the logistic regression.

## 4. Discussion

This cross-sectional study aimed to assess the prevalence of common psychiatric disorders during the COVID-19 pandemic among healthcare workers at St. Paul’s Hospital. The survey revealed a significant proportion of mental health symptoms among healthcare workers at St. Paul’s Hospital. Overall, 20.2%, 21.9%, 12.4%, and 15.5% of all respondents reported symptoms of depression, anxiety, insomnia, and psychological distress respectively. Participants involved in COVID-19 care as frontline had a higher prevalence of mental health symptoms as compared to non-frontline workers. Most participants were young (aged between 26-30 years), men, married, nurses, educated to the level of an undergraduate degree, junior in their technical title, getting monthly salary between 5,000-10,000 ETB, and have working experience of fewer than 5 years. Our study further indicates that being married was associated with experiencing severe symptoms of depression. Working in the frontline was an independent risk factor for worse mental health outcomes in all dimensions of interest. One common risk factor of the perceptions of the threat of COVID-19 in the four outcomes was thought of resigning from work due to COVID-19 (all p-values <0.01). In addition, fear of life-threatening situations once infected was associated with severe symptoms of depression. The two common protective factors of psychological protective measures in experiencing lesser symptoms of insomnia and psychological distress were satisfaction with a work-shift arrangement and logistic support (both p-values <0.01). Similarly, doctors were at lower risk of experiencing severe symptoms of anxiety and insomnia as compared to other healthcare professionals. As a result, our findings raise a concern about the psychological well-being of HCWs involved in the care and treatment of COVID-19.

Our finding is consistent with several other studies (Huang and Zhao, 2020; Que et al., 2020; Zhu *et al*., 2020) conducted during the COVID-19 pandemic. A report by Zhu et al. among 5062 HCWs in China found a significant percentage of mental health symptoms; 13.5% for depression, 24.1% for anxiety, and 29.8% for distress among healthcare workers. A study by Que et al. from China among 2285 HCWs, moderate to severe symptoms of depression, anxiety, and insomnia was 12.8%, 11.6%, and 6.8% respectively. Another recent metanalysis of thirteen studies, reported the pooled prevalence of depression to be 22.8 % and that for anxiety to be 23.2 % (Sofia *et al*. 2020). In the contrary, a report by Lai et al. in China showed symptoms of depression, anxiety, insomnia, and psychological distress among 50.4%, 44.6%, 34%, and 71.5% of HCWs respectively. This discrepancy is likely due to the differences in the methodology, where Lai et al. used lower threshold values (scores) for the diagnosis of symptoms of depression, anxiety, insomnia, and distress that overrated the prevalence.

After adjusting for confounding factors, working in a frontline position was an independent risk factor for all mental health outcomes, compared to working in a non-frontline position. Our finding is in line with other recent studies (Alkhamees *et al*., 2020; Huang and Zhao, 2020; Lai *et al*., 2020; Que *et al*, 2020; Zhu *et al*., 2020), which reported a significantly higher proportion of severe symptoms of mental illness among frontline than non-frontline HCWs. Frontline medical workers might be exposed to much more physical and mental stresses, which can contribute to their higher rates of mental problems. For example, frontline medical workers have had to be very cautious when working in COVID wards. In addition, the rapid increase of the infected patients and shortage of personal protective equipment can increase the enormous workload and psychological burden of medical workers (Cai *et al*., 2020; Xiang *et al*., 2020). Our findings revealed poor mental health among frontline medical workers. However, we did not observe a significantly higher rate of seeking help or receiving treatment for mental problems among these subjects. The phenomenon that HCWs have difficulty accepting and disclosing mental health issues is not unique to the COVID-19 outbreak (Tysser *et al*,, 2004; Fridner *et al*., 2012). Psychological distress is common among HCWs, many of whom do not seek psychological support from their colleagues, because they either think they did not need or are embarrassed to seek help and worried about confidentiality (Fridner *et al*., 2012). These findings remind that psychologists or psychotherapists should pay more attention to HCWs with mental health problems.

An interesting finding of our study was that thought of resigning from work due to COVID-19 was associated with higher risk of severe symptoms of depression, anxiety, insomnia, and distress. In contrary, being satisfied with work-shift arrangements, logistic support, and accommodation implemented by the hospital administration were the protective factors associated with lower risk of severe symptoms of mental disorders. A similar finding was recently reported by Zhu et al. from a study conducted during the earlier stage of COVID-19 pandemic in China (Zhu *et al*., 2020).

Similar to a study from China (Zhu *et al*., 2020), our finding further indicated that HCWs who were married reported more severe symptoms of depression. This might be due to increased occupational exhaustion and family responsibilities among married than unmarried HCWs. In addition, doctors, nurses, and midwives were at lower risk of experiencing severe symptoms of anxiety and insomnia than laboratory and pharmacy professionals. This could be explained by the fact that higher proportion of doctors and nurses received medical training related to COVID-19 by the hospital administration as compared to laboratory and pharmacy professionals which should be established with further studies.

The strengths of the study were the use of random sampling method and adequate number of participants. However, our study has several limitations which should be addressed in the future. First, it was a single-centre study and the findings may not be nationally representative. Second, it was a cross-sectional study and was not the best method of determining correlation and causation. Third, all the data collected were self-reported by the respondents and could be exposed to social desirability bias and more objective data can be used in future similar research.

## 5. Conclusion and Recommendation

In conclusion, the study demonstrates that a significant proportion of healthcare workers are suffering from symptoms of mental health disorders during COVID-19 pandemic at St. Paul’s Hospital. Being married and working in frontline were independently associated with a greater risk of experiencing severe symptoms of mental disorders.

Strategies to provide psychological support should be implemented by the hospital administration for the mental health of HCWs in order to control the impact of the Pandemic. Long-term surveillance should be implemented to monitor the mental health of frontline and non-frontline medical workers.

## Data Availability

The datasets generated during and/or analyzed during the current study are available from the corresponding author on reasonable request.

## Authors’ contributions

HAM and MT conceived and developed the study protocol and drafted the manuscript. All authors involved in data collection. Interpretation and final review were done by HAM, MT, EW, and TB. All authors approved the final manuscript.

## Acknowledgements

We would like to thank St. Paul’s Hospital Millennium Medical College Research Directorate for funding the study. We would like to thank all healthcare workers who, despite the increased workload during the COVID-19 crisis, have dedicated their time to answer the survey.

## References

Alkhamees, A. A., Alrashid, S.A., Alzunaydi, A.A, Almohimeed, A.S., Aljohani, M. S., 2020. The psychological impact of COVID-19 pandemic on the general population of Saudi Arabia. Comprehensive Psychiatry 102, p. 152192. doi: 10.1016/j.comppsych.2020.152192.

Fridner, A., Belkić, K., Marini, M, Sendén M. G., Schenck-Gustafsson, K., 2012. Why don’t academic physicians seek needed professional help for psychological distress□? Swiss Medical Weekly, 142, p. w13626. doi: 10.4414/smw.2012.13626.

Bohlken, J., Schömig, F., Lemke, M.R., Pumberger, M., Riedel-Heller, S. G., 2020. COVID-19 Pandemic□: Stress Experience of Healthcare Workers. Psychiat Prax. 47, pp. 190–197.

Cai, Q., Feng, H., Huang, J., Wang, M., Wang, Q., et al., 2020. The mental health of frontline and non-frontline medical workers during the coronavirus disease 2019 (COVID-19) outbreak in China: A case-control study. Journal of Affective Disorders, 275, pp. 210–215.

Creamer, M., Bell, R. and Failla, S., 2003. Psychometric properties of the Impact of Event Scale-Revised. Behaviour Research and Therapy, 41, pp. 1489–1496. doi: 10.1016/j.brat.2003.07.010.

Gelaye, B., Williams, M.A., Lemma, S., Deyessa, N., Bahretibeb, Y. et al., 2014. Validity of the Patient Health Questionnaire-9 for depression screening and diagnosis in East Africa. Psychiatry Res. 210(2). doi: 10.1016/j.psychres.2013.07.015.

Greenberg, N., Docherty, M., Gnanapragasam, S., and Wessely, S., 2020. Managing mental health challenges faced by healthcare workers during covid-19 pandemic Early support. BMJ, 368, p. m1211. doi: 10.1136/bmj.m1211.

Guan, W., Ni, Z., Hu, Y., Liang, W., Ou, C., et al., 2020. Clinical characteristics of coronavirus disease 2019 in China. New England Journal of Medicine, 382(18), pp. 1708–1720. doi: 10.1056/NEJMoa2002032.

Hanlon, C., Medhin, G., Selamu, M., Breuer, E., Worku, B., et al., 2015. Validity of brief screening questionnaires to detect depression in primary care in Ethiopia. Journal of Affective Disorders. Elsevier, 186, pp. 32–39. doi: 10.1016/j.jad.2015.07.015.

Huang, Y. and Zhao, N., 2020. Generalized anxiety disorder, depressive symptoms and sleep quality during COVID-19 outbreak in China: a web-based cross-sectional survey. Psychiatry Research journal, 288, p. 112954. doi:10.1016/j.psychres.2020.112954.

Que, J., Shi, L., Deng, J., Lui, J., Zhang, L., et al., 2020 Psychological impact of the COVID-19 pandemic on healthcare workers: a cross-sectional study in China. General Psychiatry, 33, p. e100259.

Johnson, S. U., Ulvenes p. G., Øktedalen, T., and Hoffart, A., 2019. Psychometric Properties of the General Anxiety Disorder 7-Item (GAD-7) Scale in a Heterogeneous Psychiatric Sample. Front. Psychol., 10, p. 1713. doi: 10.3389/fpsyg.2019.01713.

Lai, C., Shih, T., Ko, W., Tang, H., and Hsueh, P., 2020. Severe acute respiratory syndrome coronavirus 2 (SARS-CoV-2) and coronavirus disease-2019 (COVID-19): The epidemic and the challenges. International Journal of Antimicrobial Agents. Elsevier B.V., 55(3), p. 105924. doi: 10.1016/j.ijantimicag.2020.105924.

Lai, J., Ma, S., Wang, Y., Cai, Z., Hu, J., et al., 2020. Factors Associated With Mental Health Outcomes Among Health Care Workers Exposed to Coronavirus Disease 2019. JAMA network open, 3(3), p. e203976. doi: 10.1001/jamanetworkopen.2020.3976.

Li, S., Wang, Y., Xue, J., Zhao, N., and Zhu, T., 2020. The Impact of COVID-19 Epidemic Declaration on Psychological Consequences□: A Study on Active Weibo Users. Int. J. Environ. Res. Public Health, 17, p. 2032.

Liu, X., Kakade, M., Fuller, C. J., Fan, B., Fang, Y., Kong, J., Guan, Z., and Wu, P., 2020. Depression after exposure to stressful events: lessons learned from the severe acute respiratory syndrome epidemic. Comprehensive Psychiatry, 53, pp. 15–23. doi: 10.1016/j.comppsych.2011.02.003.

Ong, J. C., & Suh, S., 2013. Diagnostic Tools for Insomnia, in Encyclopedia of Sleep. Elsevier Inc., pp. 268–273.

Pappa, S., Ntella, V., Giannakas, T., Giannakoulis, V.G., Papoutsi, E., 2020. Prevalence of depression, anxiety, and insomnia among healthcare workers during the COVID-19 pandemic: A systematic review and meta-analysis. Brain, Behavior, and Immunity, 88, p. 901–907. doi: https://doi.org/10.1016/j.bbi.2020.05.026.

Tysser, R., Røvik J. O., Vaglum, P., Gronvold, N. T., Ekeberg, O., 2004. Help-seeking for mental health problems among young physicians: is it the most ill that seeks help□? A longitudinal and nationwide study. Soc Psychiatry Psychiatr Epidemiol., 39, pp. 989–993. doi: 10.1007/s00127-004-0831-8.

World Health Organization, 2020. Update on COVID-19. Situation report, September 18. World Health Organization.

Xiang, Y., Zhao, Y., Liu, Z., Li, X., Zhao, N., Cheun, T., and Ng, C., 2020. The COVID-19 outbreak and psychiatric hospitals in China: managing challenges through mental health service reform, Int. J. Biol. Sci., 16, pp. 1741–1744. doi: 10.7150/ijbs.45072.

Zhu, Z., Xu, S., Wang, H., Wu, J., Li, G., et al., 2020. COVID-19 in Wuhan: Immediate Psychological Impact on 5062 Health Workers. medRxiv.,1095. doi: 10.1101/2020.02.20.20025338.

